# Time trends and modifiable factors of contact tracing coverage in Geneva, Switzerland, June 2020 to February 2022

**DOI:** 10.1101/2023.03.22.23287577

**Authors:** Denis Mongin, Nils Bürgisser, the Covid-SMC Study Group, Delphine Sophie Courvoisier

**Author notes:** Membership of the Covid-SMC Study Group is provided in the supplementary material. **Correspondence**: Denis Mongin.

## Abstract

**Background:** Contact tracing has been one of the central non-pharmaceutical interventions implemented worldwide to try to control the spread of Sars-CoV-2, but its effectiveness strongly depends on its ability to detect contacts.

**Methods:** We analysed 166’892 concomitant infections occurring at the same address from June 2020 until February 2022 using an extensive operational database of SARS-CoV-2 tests in Geneva and used permutations statistics to compare the total number of secondary infections occurring at the address with those reported through contact tracing.

**Results:** Manual contact tracing captured on average 41% of the secondary infections, with variation in time from 23% during epidemic peaks to 60% during low epidemic activity. People living in wealthy neighbourhoods were less likely to report contacts (adjusted odds ratio (aOR): 1.6). People living in buildings, compared to people living in single house, were also less likely to report contacts than those living in houses, with an aOR of 1.1 to 3.1 depending on the variant, the size of the building and the presence of shops. This under-reporting of contacts in buildings decreased during periods of mandatory face masking and restriction of private gathering.

**Conclusions:** Contact tracing alone does not detect enough secondary infections to efficiently reduce the propagation of Sars-CoV-2. Public messages and outreach campaigns targeting specific populations, such as those in affluent areas, could enhance coverage. Additionally, measures like wearing face masks, improving ventilation, and implementing gathering restrictions should also be considered to reduce the number of infections occurring during interactions that may not be perceived as high risk.

## Introduction

The worldwide surge of COVID-19 in the early 2020 forced governments around the world to implement a large panel of measures, including non-pharmaceutical interventions (NPI)[1,2] to try to reduce the spread of SARS-CoV-2. These interventions ranged from lockdowns, travel restrictions, schools or public building closures, interdiction of large events to contact tracing, with variable effect on the disease transmission[1].

Among the NPI, contact tracing rapidly became a central measure to limit the transmission of the virus[3]. The idea of contact tracing is to reduce the onward spread of the virus of person who have been in contact with an infected index case, by limiting their potentially infectious contacts[4] through a reduction of social interactions and an increase of protective measures (face masks, room ventilation). The contact tracing can be manual, semi-automated after a positive test result, or based on mobile app. It can be initiated by the health authorities (state-initiated) or by the citizen (citizen-initiated). Finally, it can be exerted forward, to look for contacts of the index cases that can be infected afterward, or backward, to look for the contacts that contaminated the index case. Although theoretically effective[5], backward contact tracing had a limited use for SARS-CoV-2[6].

The efficacy of forward contact tracing is perfect if all contacts are identified (by the index case or by an app), and notified before they become contagious[7], and comply with protective measures (quarantine, face masks). In real-world settings, the true effectiveness of contact tracing for SARS-CoV-2 is estimated to range from a 63% reduction in new infections to no discernible difference[8] depending on the study and country involved. Contact tracing apps for SARS-CoV-2 are a good illustration of this phenomenon: they have received a lot of attention[9], have been developed in many places[10] and have been shown, in controlled setting, to have large potential effect[11]. Nevertheless, ecological studies obtained varying effectiveness, which can be high upon proper uptake and adherence[11] to very small, ranging from 0,1% to 11% additional infections detected by digital tracing alone[12].

There are several reasons for the relatively low effectiveness of contact tracing. First, contacts may not follow recommendations, for instance they may evade quarantine or not use protective measures such as face masks. Second, the delay in notification and the number of contacts identified[13] may limit its effectiveness, since each new contact requires a minimum amount of time to be reached[14] and not all contacts can be reached in time to apply effective measures against the spread of SARS-CoV-2. Third, there could be intentional or unintentional under-reporting. In other words, an index case may intentionally not declare contacts, or they could simply not be aware of being in contact with someone. Indeed, SARS-CoV-2 is an airborne virus[15,p.2], and multiple examples of contamination across closed space without direct encounter between index and contact cases have been reported, such as contamination through corridors[16,p.19], shared space[17], room ventilation systems[18], or even air leak through the roof[19].

The aim of this study is to estimate the number of secondary infections occurring at the same address captured by contact tracing and identify factors associated with their detection.

## Methods

To this mean, we used permutation statistics on more than 142’000 reported infections to estimate the number of secondary infections occurring at the same address. Using contact information provided by these cases, we estimate the number of infections that have been declared as contacts and assess its association with demographic and socio-economic factors.

### Data

We used all registered tests performed by persons living in the state of Geneva, Switzerland, from the Actionable Register of Geneva Outpatients and inpatients with SARS-CoV-2 (ARGOS) database[20], which is an ongoing operational COVID-19 database created by the Geneva health state agency (Geneva Directorate of Health). The register contains sociodemographic details, baseline and follow-up COVID-19 related health indicators and contact information. Due to privacy issues, these individual-level Data are available upon request at https://edc.hcuge.ch/surveys/?s=TLT9EHE93C. Response is provided within two weeks. Data are provided de-identified and thus exact address is not available. The code used to analyse the data and produce the tables and figures is available at https://gitlab.com/dmongin/scientific_articles/-/tree/main/contact_tracing.

### Study period and setting

We used data from the 01-06-2020 to 1^st^ February of 2022 having an address (3.4% of the infections did not have an address). Geneva is a mainly urban state of 511’921 inhabitants as of the last census in December 2021, with a high population density (∼13’000 hab/km^2^). It is divided geographically in 417 administrative neighbourhoods (sous-secteurs) with a median population of around a thousand persons. Each address of the dataset was geocoded using the exhaustive list of all addresses of the State of Geneva, and each neighbourhood area was associated with a socio-economic indicator provided by the centre for analysis of inequalities (hereafter the CATI-index) ranging from 0 (wealthy) to 6 (poorest). This index is then categorized in four categories, similarly to a previous study [21] (see details in supplementary material).

As the ARGOS data did not contain information about the SARS-CoV-2 variant type, we divided the study period into period of predominance of SARS-CoV-2 variants. Similarly to another study based on the same data [22], we modelled the evolution of variants based on the data provided by Covariants and the Global Initiative on Sharing Avian Influenza Data[23] in the Geneva region and defined the periods when the variant were above 50% of all variants:

– EU1 from 01-06-2020 to 05-01-2021
– Alpha from 06-01-2021 to 14-06-2021
– Delta from 15-06-2021 to 17-12-2021
– Omicron from 18-12-2021 to 01-02-2022 (mainly BA.1)

Detail about the calculations, about the different NPIs in place during these periods, and about the vaccine used in Geneva is stemming from [22] and is available in supplementary materials.

### Definition, declaration and follow up of contacts

Person testing positive for SARS-CoV-2 had the legal obligation to declare their contacts. Contacts were defined by the Swiss Confederation as persons having had an interaction with the infected person during at least 15 min at less than 1.5m, up to 48 hours before the index’ symptoms and up to 5 days after the test in absence of symptoms.

Declared contact in Geneva had the obligation to quarantine during 10 days since the implementation of contact tracing the 27^th^ of April 2020. Children below 12 years were exempted of quarantine. The 8^th^ February, 2021, it was allowed to shorten the quarantine at day 7 by providing a negative nasopharyngeal or oropharyngeal SARS-CoV-2 PCR test. The quarantine was shortened to 7 days the 31th of December 2021, and to 5 days the 12^th^ of January 2022. By the end of 2021, vaccinated persons (at least 2 doses of mRNA vaccination Moderna mRNA- 1273 or Pfizer BNT162b2 or one dose of Janssen Ad26.COV2-S vaccine) or persons with a positive test during the last 4 month did not have the obligation to quarantine after a contact with an infected index. Since October 2020, health professionals were allowed to work even if they were contacts of an infected person. They were however systematically tested by their institution and if they tested positive, they had to isolate for at least 48 hours. They could go back to work after 48 hours if they show only mild symptoms and no fever, while pursuing barrier measure during at least 7 days. A graphical timeline is provided in supplementary material to summarize these changes.

In January 2021, an anthropologist was hired by the state COVID unit, who trained the tracing team in motivational interviewing and performed field study to understand the barriers and facilitating factor to declare one’s contact.

Contact information was initially collected by personal telephone interviewing of the index case (February 2020 to end of April 2020). From May 2020, index cases had the possibility to provide their contacts names and phone through an online form, and information was completed when the index case was called. Contacts were sent a message telling them they were contact and should quarantine, and were then contacted by phone. Additionally, an online form was implemented at the end of September 2020 to support the phone calls, where the contacts could complete the required information themselves. From mid December 2021, the phone calls could not be maintained due to the high number of cases, therefore contact information was only obtained from the online forms.

### Outcomes: secondary infections occurring at the same address and absence of reporting

Coverage of contact tracing could be roughly estimated by divided the number of infections captured by the contact tracing by the total number of infections recorded. Although simple, this method has two caveats. First, Geneva is a region sharing its border with France with a population doubling every day because of workers commuting between the two countries. France having its own contact tracing system, this makes the number of secondary infections of French citizen working in Geneva difficult to estimate. Secondly, this raw calculation does not allow to consider modifiable factors such as socio-economic condition or living conditions. We thus decided to restrict our study to secondary infection occurring at the same address in Geneva, which will be our primary outcome.

To estimate this number, we first calculated concurrent infections of two persons living at the same address and having a positive COVID-19 result less than 10 days apart. The date associated with the concurrent infection was the middle date between the two test results. We then used the exhaustive list of declared contacts to define the binary variable “absence of reporting” as being 0 if the concurrent infection was captured by the contact tracing and 1 otherwise. We considered the possibility that the concurrent infection was declared by the index or by the contact and did not restrict to any specific form of relation type declared by the index.

Concurrent infections capture both infections that are related to the fact of living at the same address (secondary infections), and infections that are occurring by chance at the same address (concomitant infections: i.e. two person living at the same address can be infected 10 days apart by other persons anywhere else).

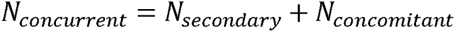

In order to determine the number of secondary infections, one must estimate the number of concomitant infections, corresponding to a null hypothesis that there are no excess infections due to the fact of living at the same address (H0: N*N_secondary_* = 0). In other words, under this null hypothesis, concurrent infections occurring at the same address are due to chance only (*N_concurrent_* = *N_concomitant_*). Permutation techniques can be used to estimate the frequency of concurrent infections under the null hypothesis [24]. It consists in permuting randomly without replacement each person’s addresses to then compute the number of concurrent infections at the same address. One obtains in this situation what we called concomitant infection, that is the number of infections occurring at the same address only by chance (because the addresses were permuted). We can then estimate the number of secondary infections occurring at the living address as the difference between the raw number of concurrent infections at a given address, obtained in the ARGOS register and the ones obtained by permutation. We performed 1000 permutations and operationalized the estimation of secondary infections as the median value of the difference obtained. To account for potential confounding, the addresses were permuted within each neighbourhood and within each type of building. Permuting within each type of neighbourhood allows to avoid confounding caused by the socio-economic condition of the neighbourhood or by shared services, such as schools, grocery stores, and some of the public transportations. Permuting within each building type allows to avoid confounding caused by the association between concomitant infections and the size of the building. Indeed, the probability to have a concurrent infection by chance for two persons living at the same address is higher in a large building than in a small house.

### Statistical analysis

Confidence interval of estimation of secondary infections occurring at the same address was operationalized as the 2.5% and 97.5% quantile of the difference between the number of concurrent infections at a given address and the concomitant infections obtained by permutation. This analysis was stratified by variants.

To examine the association of gender, vaccination, living characteristics and socio-economic characteristics with potential under-reporting of contacts, we applied to concurrent infections calculated on the initial dataset and on each permutations a generalized linear model using the absence of reporting as outcome, with CATI index, type of building, number of people living at the address, immune status of the two persons of the concurrent infection dyads and their gender as covariate. The immune status was recalculated for each permutation at the date corresponding to the corresponding concurrent infections. The final estimates of the model were given by the median and 2.5% 97.5% quantile of the differences between the coefficients obtained on the raw dataset and the ones obtained on each of the permuted datasets.

All analysis were performed using R.4.1.0, and the high performance computing facility “Baobab” of the University of Geneva.

### Covariates

The immune status was operationalized as fully vaccinated if both persons were vaccinated with at least one dose, mixed if one of the two was vaccinated with at least one dose, and not vaccinated if none was vaccinated. The type of vaccines considered is described in supplementary material. The immune status was calculated at the date of secondary infection. Gender was operationalized as men if both persons were men, women if both were women, and mixed otherwise.

As contact tracing coverage may be influenced by the number of social interactions and the environment of the encounter, we categorized the type of building considering both the population living at the address and the type of building. We thus decided to consider separately addresses where 2 or less people lived, as they were less kind to have social interaction with people living with them, from houses or buildings and we distinguished the presence of shops at the address or not. The type of building was therefore operationalized in 6 categories (detailed explanation in supplementary material):

- Building at the address with up to 2 inhabitants (houses with isolated persons)
- Building with more than 2 inhabitants:

`o` Houses (family houses)
`o` Building with no shops and less than 40 inhabitants
`o` Building with no shops and more than 40 inhabitants
`o` Building with shops and less than 40 inhabitants
`o` Building with shops and more than 40 inhabitants

Of note, in the Geneva region, buildings in the city often have shops on the ground floor, compared to buildings in more rural areas.

### Sensitivity analysis

The delay between two positive tests at the same address used to define concurrent infections has been set in the main analysis as twice 5 days, the mean incubation period of early variants [25]. As this delay could influence our results, we performed two sensitivity analysis by defining concurrent infections with a shorter delay of 6 days (twice three days) or 14 days (twice 7 days) between the two positive tests.

## Results

Over the period considered, 25’297 addresses had at least two persons with a positive test result less than 10 days apart (i.e. at least one concurrent infection, table 1). The median number of concurrent infection dyads at these addresses was 3 (Inter Quartile Range IQR: 1-6), though it was lowest during the alpha wave (1 IQR: 1-3) and highest during the omicron wave (3 IQR: 1-10). The addresses were mainly situated in the wealthiest (37%) and poorest areas (29%), and concerned a median amount of 29 persons, with no notable change across time. The main type of building were buildings with no shops and less than 40 inhabitants (32%), followed by buildings with shops and more than 40 inhabitants, buildings with shops and less than 40 inhabitants (17%), family houses (16%), building with shops and more than 40 inhabitants (13%) and houses with maximum 2 persons (3%).

**Table 1:** characteristics of the addresses at which at least one concurrent infection occurred, for the overall period (overall) and stratified by variant. CATI score is the socio-economic index of the neighbourhood, with 0 the wealthiest and 6 the poorest.

|  | **Overall** | **EU1** | **alpha** | **delta** | **omicron** |
| --- | --- | --- | --- | --- | --- |
| Number of addresses | 25'196 | 6'596 | 2'805 | 5'007 | 10'788 |
| Median number of concurrent infections per address | 3.00<br>IQR: 1.00-6.00 | 2.00 IQR: 1.00, 4.00 | 1.00 IQR: 1.00-3.00 | 2.00 IQR: 1.00-4.00] | 3.00 IQR: 1.00-10.00 |
| CATI score |  |  |  |  |  |
| 0 | 9'238 (37.0%) | 2'334 (35.6%) | 936 (33.6%) | 1'881 (38.0%) | 4'087 (38.3%) |
| 1 | 3'939 (15.8%) | 992 (15.1%) | 476 (17.1%) | 773 (15.6%) | 1'698 (15.9%) |
| 2-3 | 4'419 (17.7%) | 1'206 (18.4%) | 496 (17.8%) | 844 (17.0%) | 1'873 (17.6%) |
| 4-6 | 7'360 (29.5%) | 2'027 (30.9%) | 874 (31.4%) | 1'456 (29.4%) | 3003 (28.2%) |
| Building type |  |  |  |  |  |
| Family houses (refence) | 4010 (15.9%) | 873 (13.2%) | 397 (14.1%) | 838 (16.7%) | 1902 (17.6%) |
| House with isolated persons | 726 (2.9%) | 250 (3.8%) | 79 (2.8%) | 108 (2.1%) | 289 (2.7%) |
| Building without shops less than 40 inhabitants | 8076 (31.9%) | 2021 (30.5%) | 826 (29.4%) | 1548 (30.8%) | 3681 (34.0%) |
| Building without shops more than 40 inhabitants | 5029 (19.9%) | 1444 (21.8%) | 687 (24.4%) | 1075 (21.4%) | 1823 (16.8%) |
| Building with shops less than 40 inhabitants | 4256 (16.8%) | 1142 (17.3%) | 406 (14.4%) | 757 (15.0%) | 1951 (18.0%) |
| Building with shops more than 40 inhabitants | 3200 (12.6%) | 888 (13.4%) | 417 (14.8%) | 705 (14.0%) | 1190 (11.0%) |

### Excess concurrent infections (include state versus neighbourhood baseline?)

During the period of interest, 166’892 raw concurrent infections occurred (see table 2). The null hypothesis estimation yielded 117’617 CI: 116’363-118’945 concurrent infections. The estimated excess number of concurrent infections occurring at the same address is then 49’275 CI:47’947-50’529.

**Table 2:** Number of persons infected at the same address 10 days apart from each other, in the real data base, when permuting addresses on the state level (state base permutation), or on the neighbourhood level (neighbourhood base permutation). Estimations for permutation are the median of 1000 permutations given with their percentile confidence intervals (2.5% - 97.5% range).

|  | **total** | **EU1** | **alpha** | **delta** | **omicron** |
| --- | --- | --- | --- | --- | --- |
| Number of raw concurrent infections | 166'892 | 38'562 | 9'551 | 19'382 | 99'397 |
| Number of concurrent infections in permutations | 117'617 CI:<br>118'945-116'363 | 22'722 CI:<br>23'041-22'412 | 2'484 CI: 2'573-2'398 | 9'499 CI: 9'714-9'306 | 82'912 CI: 83'617-82'247 |
| Estimated contagions at the address | 49'275 CI: 47'947,<br>50'529 | 15'840 CI:<br>15'521-16'150 | 7'067 CI: 6'978-7'153 | 9'883 CI: 9'668-10'076 | 16'485 CI: 15'780-17'150 |
| Number of contacts declared living at the same address | 20'990 | 5'341 | 3'687 | 5'085 | 6'877 |
| percentage of contagions declared | 42.6% CI: 41.5-43.8 | 33.7% CI: 33.1-34.4 | 52.2% CI: 51.5-52.8 | 51.5% 50.5-52.6 | 41.7% CI: 40.1-43.6 |

### Proportion of infections reported through contact tracing

The 20’990 declared contact, living at the same address than their index case and who became positive less than 10 days apart following the index case test result, accounted for 42.6% CI:41.5-43.8 of the estimated address concurrent infections. This percentage was at its lowest during the EU1 wave with 33.7% CI: 33.1-34.4, rose above 50% for alpha and delta wave (52.2% CI: 51.5-52.8 and 51.4% CI: 50.4-52.6 respectively), and decreased to 41.8% CI: 40.0-43.6 during the omicron wave.

The monthly evolution of this percentage of infections captured by contact tracing fluctuated between 67% and 23% (figure 1), and tended to be lower when the number of COVID-19 cases was high. The lowest values of contact reported were observed during the two periods reaching more than 10’000 COVID-19 cases per month (the peak of EU1 wave, and the end of delta/start of omicron wave). Of note the strongest increase of the rate of contacts reported was in January 2021, from 23% to 50%.

**Figure 1:**
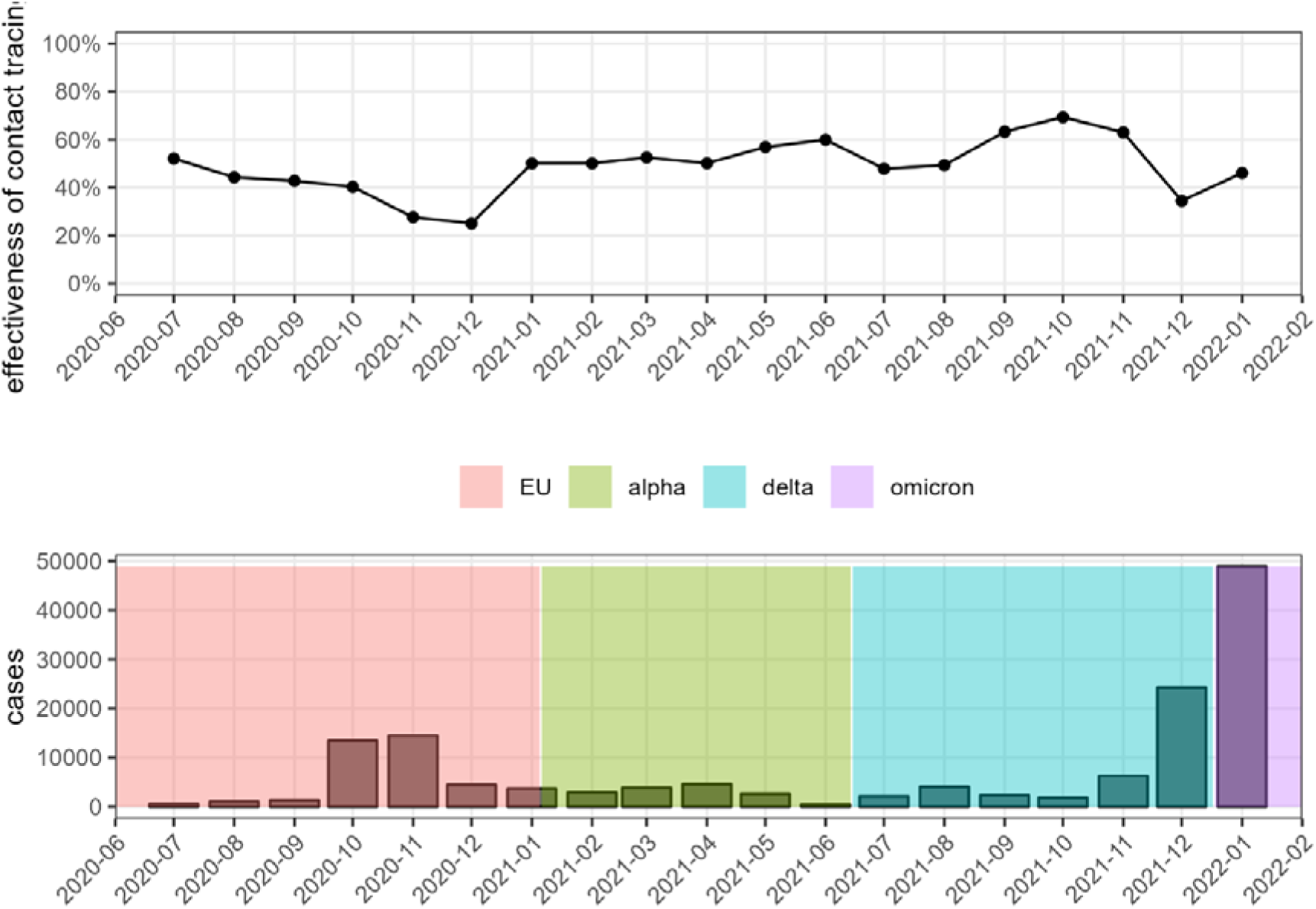
Monthly time evolution of the median percentage of address co-infection captured by the contact tracing system (upper panel), compared to the monthly evolution of the number of cases in Geneva (lower panel), for the four periods considered in the present analysis.

### Determinants of absence of reporting

When compared to the adult age category (17 to 65 years old), a contact child (age under 17) tended to have more chance to be under-reported during early variants. During the EU1 wave, an index younger than 17, especially during the omicron wave, had less chance to have its contact reported (OR 1.27 CI: 1.07-1.56).

The socio-economic status of the neighbourhood had a strong dose-response association with under-reporting: persons from the poorest neighbourhood were less likely to under-declare their contacts, with an odd ratio reaching OR 0.59 CI: 0.47-0.72 for the most disadvantaged neighbourhood during omicron (table 3).

**Table 3:**
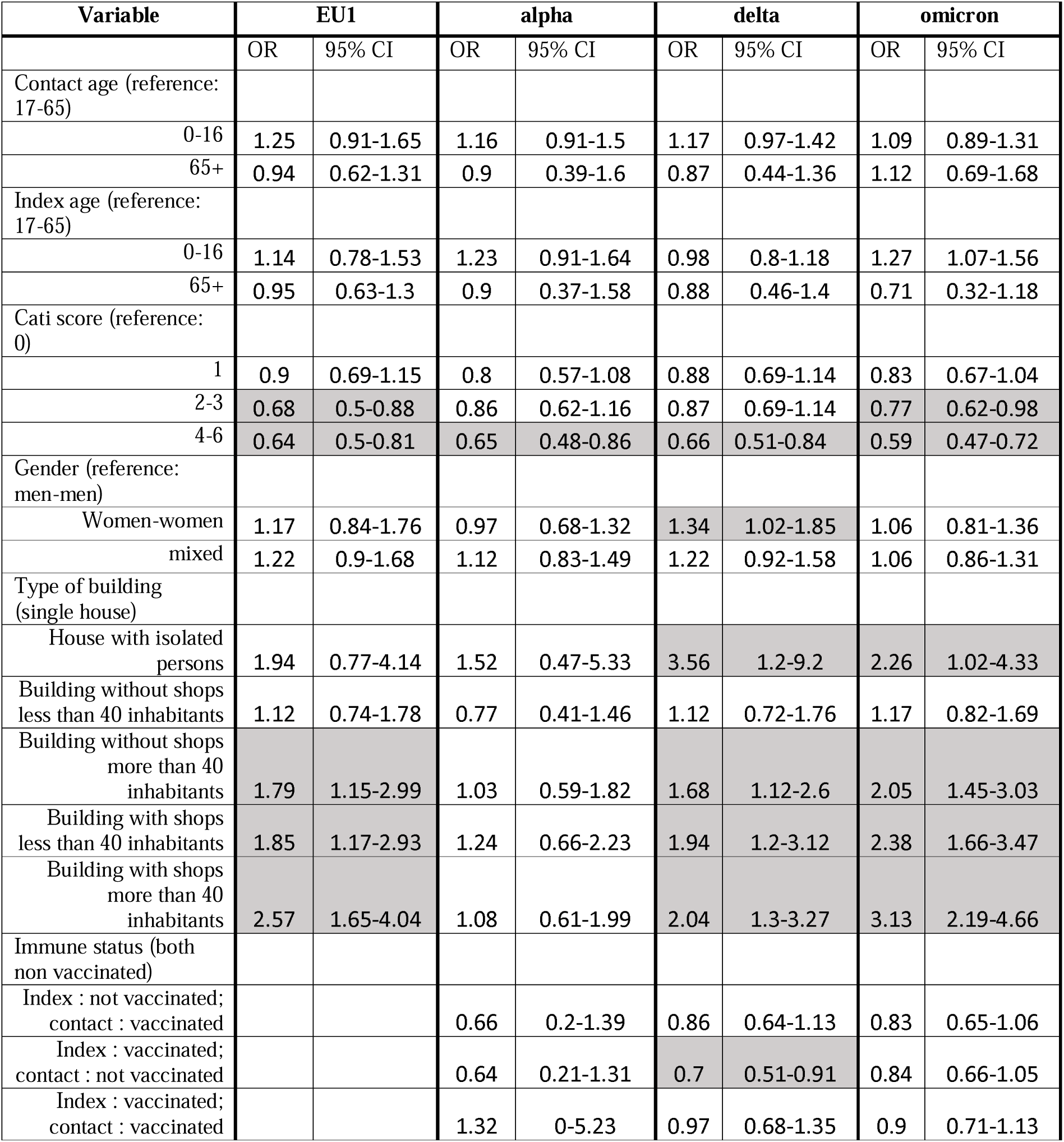
Odds ratio (OR) with their associated confidence interval of the multivariable generalized model of under-reporting of the secondary infection. OR with and IR not encompassing 1 are grey shaded.

| Variable | EU1 |  | alpha |  | delta |  | omicron |  |
| --- | --- | --- | --- | --- | --- | --- | --- | --- |
|  | OR | 95% CI | OR | 95% CI | OR | 95% CI | OR | 95% CI |
| Contact age (reference: 17-65) |  |  |  |  |  |  |  |  |
| 0-16 | 1.25 | 0.91-1.65 | 1.16 | 0.91-1.5 | 1.17 | 0.97-1.42 | 1.09 | 0.89-1.31 |
| 65+ | 0.94 | 0.62-1.31 | 0.9 | 0.39-1.6 | 0.87 | 0.44-1.36 | 1.12 | 0.69-1.68 |
| Index age (reference: 17-65) |  |  |  |  |  |  |  |  |
| 0-16 | 1.14 | 0.78-1.53 | 1.23 | 0.91-1.64 | 0.98 | 0.8-1.18 | 1.27 | 1.07-1.56 |
| 65+ | 0.95 | 0.63-1.3 | 0.9 | 0.37-1.58 | 0.88 | 0.46-1.4 | 0.71 | 0.32-1.18 |
| Cati score (reference: 0) |  |  |  |  |  |  |  |  |
| 1 | 0.9 | 0.69-1.15 | 0.8 | 0.57-1.08 | 0.88 | 0.69-1.14 | 0.83 | 0.67-1.04 |
| 2-3 | 0.68 | 0.5-0.88 | 0.86 | 0.62-1.16 | 0.87 | 0.69-1.14 | 0.77 | 0.62-0.98 |
| 4-6 | 0.64 | 0.5-0.81 | 0.65 | 0.48-0.86 | 0.66 | 0.51-0.84 | 0.59 | 0.47-0.72 |
| Gender (reference: men-men) |  |  |  |  |  |  |  |  |
| Women-women | 1.17 | 0.84-1.76 | 0.97 | 0.68-1.32 | 1.34 | 1.02-1.85 | 1.06 | 0.81-1.36 |
| mixed | 1.22 | 0.9-1.68 | 1.12 | 0.83-1.49 | 1.22 | 0.92-1.58 | 1.06 | 0.86-1.31 |
| Type of building (single house) |  |  |  |  |  |  |  |  |
| House with isolated persons | 1.94 | 0.77-4.14 | 1.52 | 0.47-5.33 | 3.56 | 1.2-9.2 | 2.26 | 1.02-4.33 |
| Building without shops less than 40 inhabitants | 1.12 | 0.74-1.78 | 0.77 | 0.41-1.46 | 1.12 | 0.72-1.76 | 1.17 | 0.82-1.69 |
| Building without shops more than 40 inhabitants | 1.79 | 1.15-2.99 | 1.03 | 0.59-1.82 | 1.68 | 1.12-2.6 | 2.05 | 1.45-3.03 |
| Building with shops less than 40 inhabitants | 1.85 | 1.17-2.93 | 1.24 | 0.66-2.23 | 1.94 | 1.2-3.12 | 2.38 | 1.66-3.47 |
| Building with shops more than 40 inhabitants | 2.57 | 1.65-4.04 | 1.08 | 0.61-1.99 | 2.04 | 1.3-3.27 | 3.13 | 2.19-4.66 |
| Immune status (both non vaccinated) |  |  |  |  |  |  |  |  |
| Index : not vaccinated; contact : vaccinated |  |  | 0.66 | 0.2-1.39 | 0.86 | 0.64-1.13 | 0.83 | 0.65-1.06 |
| Index : vaccinated; contact : not vaccinated |  |  | 0.64 | 0.21-1.31 | 0.7 | 0.51-0.91 | 0.84 | 0.66-1.05 |
| Index : vaccinated; contact : vaccinated |  |  | 1.32 | 0-5.23 | 0.97 | 0.68-1.35 | 0.9 | 0.71-1.13 |

The type of building had significant effects during the EU1, delta and omicron wave with an effect that tended to increase in time, but no significant effect during the alpha wave. When compared to family houses, under- reporting increased with the number of inhabitants in buildings and with the presence of shops. In detail, we observed no significant effect for building without shops and less than 40 inhabitants. However, we found an odds ratio (OR) for not reporting contact from OR 1.79 CI: 1.15-2.99 to OR 2.06 CI: 1.46-3.03 during EU1 and omicron respectively for building without shops with more than 40 inhabitants and OR 1.85 CI: 1.17-2.93 to OR 2.38 CI: 1.66-3.47 during EU1 and omicron respectively for building with shops and less than 40 inhabitants. The highest OR was found during EU1 and omicron for building with shops and more than 40 inhabitants (OR 2.57 CI: 1.65-4.04 to OR 3.13 CI: 2.19-4.66 respectively). Houses with isolated persons had significant difference of reporting during the delta and omicron period, but with a very large confidence interval (OR 3.56 CI: 1.20-9.20 and OR 2.26 CI: 1.02-4.33 during delta and omicron waves).

Being vaccinated favoured the declaration of contacts only when one of the two persons implied was vaccinated, the effect reaching significance during the delta wave when the index was vaccinated and the contact was not (OR 0.70 CI: 0.51-0.91) and during omicron for the opposite case (OR 0.84 CI: 0.66-1.05).

### Sensitivity analysis

Considering 6 days between two positive tests at the same address to define concurrent infections yielded a global contact coverage of 40.1% CI: 39.3-41.1 (48’895 estimated secondary infections occurring at the address CI: 47’300-50’400, for 21’733 reported contacts), while considering 14 days yielded 44.4% CI: 43.1-45.9 of contact coverage (44’875 estimated secondary infections occurring at the address CI: 43’861-45’859, for 18’017 reported contacts). The determinant of absence of reporting provided odds ratio very similar to those of the main analysis. Detailed results for the two sensitivity analysis can be found in the supplementary materials.

## Discussion

The complete database of COVID-19 infections occurring in Geneva over a period of almost 2 years allowed us to estimate the capacity of contact tracing to capture infectious contacts occurring at the same living address. In this study, on average, contact tracing allowed to detect 41% of the actual infections occurring at the living address. This percentage varied in time and was lower during the winter wave of 2020 and at the beginning of the Omicron wave. The principal determinants of absence of reporting of contacts were living in a wealthy socio-economic neighbourhood, younger age, and living in populated building with shops. Mixed vaccination status (one vaccinated, the other not) was associated with better reporting.

Contact tracing can have a sustained meaningful effect on disease transmission only for variants with relatively low effective reproduction numbers, as long as the coverage of contact is high and the delay of notification of the contacts stays short. Indeed simulation studies with isolation of cases only [14] showed that detecting only 40% of the contacts allows to control more than 80% of outbreaks for low reproductive numbers, but fails to control more than 10% of them if the effective reproduction number is of 3.5. For such reproduction number, controlling more than 80% of the outbreaks would require a contact coverage of almost 90%. Modelling studies [7,26] considering low basic reproduction numbers estimated that the effect of contact tracing started to have a real impact on the reproduction number if more than 50% of contacts were reached. Other studies[27] showed that reducing the contact tracing coverage from 80% to 40% multiply at least per 2 the probability of a large outbreak even with few cases. Given that the basic reproduction number of variants alpha, delta and omicron is above 3 and close to 8 for the latter[28,29], and that the proportion of contacts traced decreased during high viral activity periods, the impact of the manual contact tracing on the spread of COVID-19 may have been rather limited after the first two waves [30].

Several mechanisms can contribute to low coverage of contact tracing. The first one is the saturation of the contact tracing capacities, due to limited number of personnel and resources required to perform the contact tracing. A second one is intentional under-reporting, encompassing contacts not declared to avoid quarantine measures [31], but also contacts not declared because they were exempted of quarantine such as health professional, vaccinated persons or children below 12 years. A last mechanism could be non-intentional unreported contacts, which are infectious relation that are not perceived as such, such as using the elevator after someone who is infected, crossing an infected neighbor at the shop down the building, being infected across the corridor[16,p.19], etc.

Our study yield indications of these three mechanisms. The effect of the contact-tracing capacities is evidenced in our study by the decrease of the percentage of contacts reported during periods with high number of infected cases declared, reaching value as low as 20%. The increase of contact reported in January 2021 seems to correspond with the implementation of guidelines within the state COVID-19 unit to encourage the infected persons to declare their contacts during the phone interviews. Also during this month, the following measures were implemented from the 18^th^ of January 2021, and then gradually relaxed between May and June 2021: mandatory remote work if possible, mandatory face masking at work and in shops, closure of shop selling non consumer staple, and restriction of public and private gathering to 5 persons maximum. These public health measures could have helped people recognize any possible contacts and declare them accordingly.

The findings of this study suggest intentional under-reporting. For instance, the higher tendency to under-report children before the omicron wave is consistent with the exemption of kids from quarantine at the beginning of the pandemics in Geneva. Erroneous public health messages at the beginning of the pandemic could have helped foster underreporting in children. Indeed, during the first wave in April 2020, a statement from the Head of Communicable Diseases Divison at the Federal Office of Public Health in Switzerland, widely reported in news outlets, stated that infection in children was very unlikely and that transmission among children was close to none. A second clear indicator of the behavioral part of under-reporting lies in the higher chance of not declaring contacts in wealthy neighborhoods. This result may stem from the fact that persons living in wealthy neighbourhoods may have jobs allowing to remote work, and have therefore a lower need of official quarantine certificates. It is also in line with observed tendency of high social class individuals to exhibit higher unethical decision-making tendencies or higher tendency to break the law[32,33]. The third indicator is the higher reporting of contacts when one of the index or contact is vaccinated, which may be due to the perception that contact tracing is more useful, or reflect a better compliance to national guidelines [34].

Finally, the effect of building type on the propensity to report contacts supports the existence of infectious contacts between persons that are not identified as such. Indeed, the fact that under-reporting was higher in buildings than in family houses, especially during the Omicron wave, suggests the occurrence of unperceived contagions in the common areas (i.e. contagion where the index case did not perceive the contact as at risk of contagion), which are more numerous and common for buildings than for houses. Under-reporting was higher in building with more inhabitants and in buildings with shops, indicating that part of these un-noticed contacts may happen in shared social places. This type of shared space, such as elevator, corridors, stairs or entrance halls, do not allow proper physical distancing and are often poorly ventilated, thus allowing the concentration of potentially infectious aerosol. It could also be due to the fact that buildings with shops are more frequent in urban area, with higher population density. The absence of effect of the building type on under-reporting of contacts during the alpha wave (06-01-2021 to 14-06-2021) can be explained by the health policies implemented during that period. This finding indicates that these public health policies reduced the amount of unperceived contagions.

There are several limitations in this study. First, due to our analysis design, we restricted the study to infection occurring at the address. As a consequence, tests results without addresses (3.4%) were removed from our analysis, leading to potential selection bias. Another potential limitation of the use of addresses is that part of the reported addresses may not correspond to the actual place of living. This type of misclassification bias may underestimate the number of secondary infections (ie. bias toward the null). Also, our analysis does not consider infectious contacts occurring at other places, and similar analysis performed at the working place or in different settings could be of interest. Second, the use of aggregated socio-economic indicator at the neighbourhood level could cause ecological fallacy, where the effect observed is caused by a variable at the person level. Third, as our study is based on positive registered test, it ignores all COVID-19 positive persons who performed only self-test or did not test (because they did not want to, or because they were asymptomatic). Although most self-tests were secondarily confirmed by an officially registered testing, the real coverage of all secondary infections occurring at the address is likely lower than the one reported in the present study. Finally, as with every observational study, we cannot rule out residual confounding in the multivariable analysis, although the rich register data allowed adjusting for most of the important factors.

Nevertheless, this study offers a solid estimation of the proportion of reported infectious contacts at a given address using an extensive operational register of all SARS-CoV-2 tests performed in the state of Geneva during a period covering four predominant variants. The analysis based on permutation tests at the neighbourhood level allowed to minimize the amount of contaminations occurring at other places such as schools, grocery shops or public transportation, thus providing insights into the systemic, behavioural and living factors influencing the report of contacts. The sensitivity analysis conducted show the robustness of our results.

The overall contact coverage estimated in our study and its decrease during high epidemic activity periods indicates that contact tracing alone cannot mitigate late variants of SARS-CoV-2. Contact tracing coverage could be improved by social outreach targeting population such as those living in wealthy neighbourhoods. To further reduce the propagation of SarS-CoV-2, public health authorities should consider additional non pharmaceutical intervention aiming to avoid unperceived contagions, such as face mask wearing, air cleaning or gathering restrictions.

## Statements

### Ethical statement

Research received the agreement of the Cantonal Ethic Committee of Geneva (CCER protocol 2020-01273). Individuals who refused to share their data were removed from the analysis.

### Data availability

The *de-identified* database *underlying this article will be shared on reasonable request* using the <u>form</u> (https://edc.hcuge.ch/surveys/?s=TLT9EHE93C). The code used for the analysis is available at the following repository: https://gitlab.com/dmongin/scientific_articles/-/tree/main/contact_tracing.

## Acknowledgements

We thank the Geneva Directorate of Health for collecting and providing the data.

## Authors’ contributions

Denis Mongin performed the data curation, participated in the analysis conception, performed the statistical analysis, created the data visualisation, designed and wrote the article; Nils Bürgisser participated to the literature review and to the writing of the article, participated to the result interpretation and revised critically the article; Delphine S. Courvoisier acquired the financial support for the project, conceptualised the analysis, participated in the data interpretation and revised critically the article. Denis Mongin, Guillaume Schimmel, Adriana Uribe and Delphine Courvoisier had full access to the dataset. Denis Mongin and Delphine Courvoisier verified the data. All authors were responsible for the decision to submit for publication.

Lucienne Da Silva Mora, Diem-Lan Vu, Lena Després, Rachel Dudouit, Béatrice Hirsch, Barbara Müller, Charlotte Roux, Géraldine Duc, Caroline Zahnd, Adriana Uribe Caparros, Guillaume Schimmel, Jean-Luc Falcone, Nuno M Silva, Thomas Goeury, Christophe Charpilloz, Silas Adamou, Pauline Brindel, Roberta Petrucci, Andrea Allgöwer, Abdel Kadjangaba, Christopher Abo Loha, Emilie Macher, Marc Vassant, Nadia Donnat, Philippe Pittet, Dominique Joubert, Samia Carballido, Ariane Germain, Sophie Bontemps, Elisabeth Delaporte, Camille Genecand, Aliki Metsini, Valérie Creac’h, Virginie Calatraba, Laura Flüeli, Hippolyte Piccard, Dan Lebowitz, Aglaé Tardin, Simon Regard, as members of the Covid-SMC Study Group, participated in the collection of data, study design and revised critically the article.

