## Supplementary material for "Time trends and modifiable factors of contact tracing coverage in Geneva, Switzerland, June 2020 to February 2022"

Supplementary materials

Table des matières

### Covid-SMC Study Group

The COVID-SMC study group is composed of Lucienne Da Silva Mora, Lena Després, Rachel Dudouit, Béatrice Hirsch, Barbara Müller, Charlotte Roux, Géraldine Duc, Caroline Zahnd, Adriana Uribe Caparros, Guillaume Schimmel, Jean-Luc Falcone, Nuno M Silva, Thomas Goeury, Christophe Charpilloz, Silas Adamou, Pauline Brindel, Roberta Petrucci, Andrea Allgöwer, Abdel Kadjangaba, Christopher Abo Loha, Emilie Macher, Marc Vassant, Nadia Donnat, Philippe Pittet, Dominique Joubert, Samia Carballido, Ariane Germain, Sophie Bontemps, Elisabeth Delaporte, Camille Genecand, Aliki Metsini, Valérie Creac’h, Virginie Calatraba, Laura Flüeli, Hippolyte Piccard, Dan Lebowitz, Aglaé Tardin, and Simon Regard.

### Methods

#### Covariates

##### Building types

In the original dataset, we used the official classification used in Switzerland, which includes the following categories:

- “Individual houses”
- “Houses with several livings” (a category which also include buildings, but without annex usage such as shops or others)
- “Building with annex usage”
- “Building with partial use for living” (e.g. schools where there is a janitor living)
- “Building without living”. This category was not considered and absent from our dataset of test results.

The dataset of all address of Geneva provided by the state of Geneva included also the number of housing at the address, and the number of people living at the address. There were ~10% missing for the official building categories and the number of housing at the address. The missing were imputed as follows:

- number of housing: as the rounded mean of the number of housing of the 4 closest address with non-missing information
- official building categories: as the predominant category in the 10 closest sites with non-missing information.

Our building category was then operationalized as follow:

- house with isolated persons: any building with less or equal than 2 persons
- Houses (family houses): “Individual houses”, or “Houses with several livings” with less or equal than 2 housings
- Building with no shops and less than 40 inhabitants: “Houses with several livings” with more than 2 housings or building with partial use for living and less than 40 people living at the address
- Building with no shops and more than 40 inhabitants: “Houses with several livings” with more than 2 housings or building with partial use for living and less than 40 people living at the address
- Building with shops and less than 40 inhabitants: “Building with annex usage” with between 2 and 40 persons living at the address
- Building with shops and more than 40 inhabitants: “Building with annex usage” with more than 40 persons living at the address

##### CATI index

Socio-economic vulnerability was assessed using a neighbourhood socio-economic vulnerability index defined by the Center for Territorial Analysis of Inequalities (CATI-GE). The state of Geneva provided for each area 6 variables corresponding to different aspects of vulnerability: the proportion of household perceiving a housing allowance, the median income of households, the share of low income, the share of students coming from a modest family, the share of active persons registered to the unemployment office, and the share of persons perceiving social subsidies (e.g., disability subsidies, health insurance subsidies, or supplementary pension benefits). Table 1 summarizes these variables and indicates the threshold used to determine if the variable contribute the socio-economic vulnerability.

| **Variable** | **Threshold defining a low socio-economic level in the geographical area** |
| --- | --- |
| Proportion of housing allowance | Above the fourth quartile of the overall proportion of housing allowance |
| Median income | Below the first quartile of the overall distribution of median income |
| share of low income | Above the fourth quartile of the overall share of low income |
| share of students coming from a modest family | Above the fourth quartile of the overall share of student coming from a modest family |
| share of active persons registered to the unemployment office | Above the fourth quartile of the overall share of active persons registered to the unemployment office |
| share of person perceiving social subsidies | Above the fourth quartile of the overall person perceiving social subsidies |

Table 1 Composition of the neighbourhood socio-economic vulnerability index (NSVI). Each variable is computed for the geographical area.

The CATI index for each area was operationalized as the sum of the six dichotomized variables (1 low socio-economic level, 0 non-low socio-economic level), resulting in a score ranging from 0 to 6. In order to have similar sizes of group, we defined four groups: the reference wealthy group of area with an CATI index equal to 0, slightly socio-economic vulnerable areas with an CATI index of 1, moderately socio-economic vulnerable areas with an CATI index between 2 and 3, and highly socio-economic vulnerable areas with a CATI index higher than 3.

#### Definition, declaration and follow up of contacts.

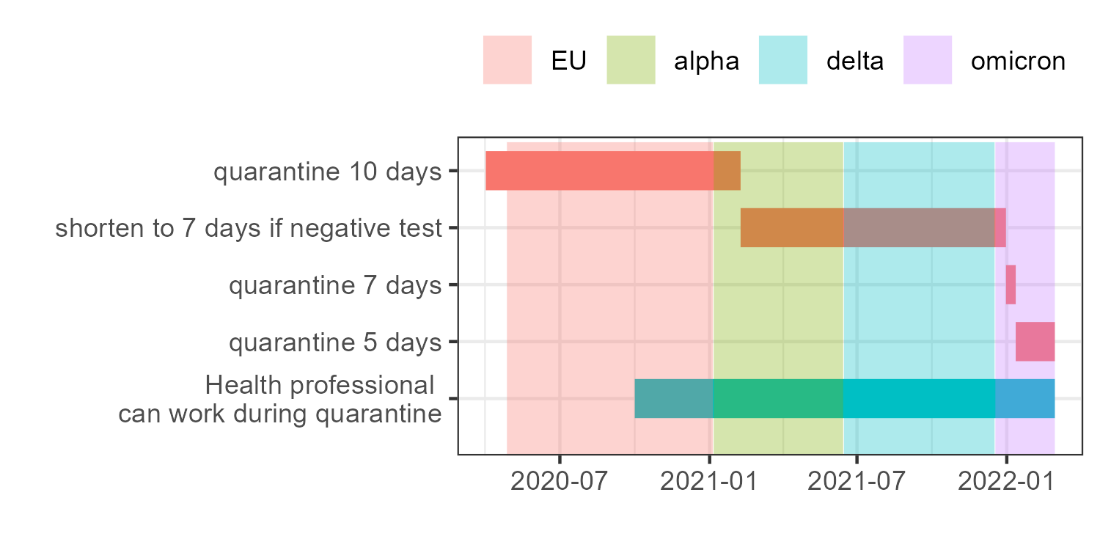

#### SARS-CoV-2 variants

The data used to determine the period of Variant predominance are provided by GISEAD and covariant.org, and concern the regions 1 of Switzerland, encompassing Geneva, Valais and Vaud cantons (<https://covariants.org/per-country?region=Switzerland>). The data were available at the following github repository <https://github.com/hodcroftlab/covariants/blob/master/cluster_tables/SwissClusters_data.json> and are provided in the gitlab repository of this article. After selecting the data for the region1 of Switzerland and the four variants of interest in our study, we modelled the VoC evolution as a rising and decreasing sigmoid in time. This approach yields the estimations presented in the graph below (estimations are solid lines, points are the experimental data)

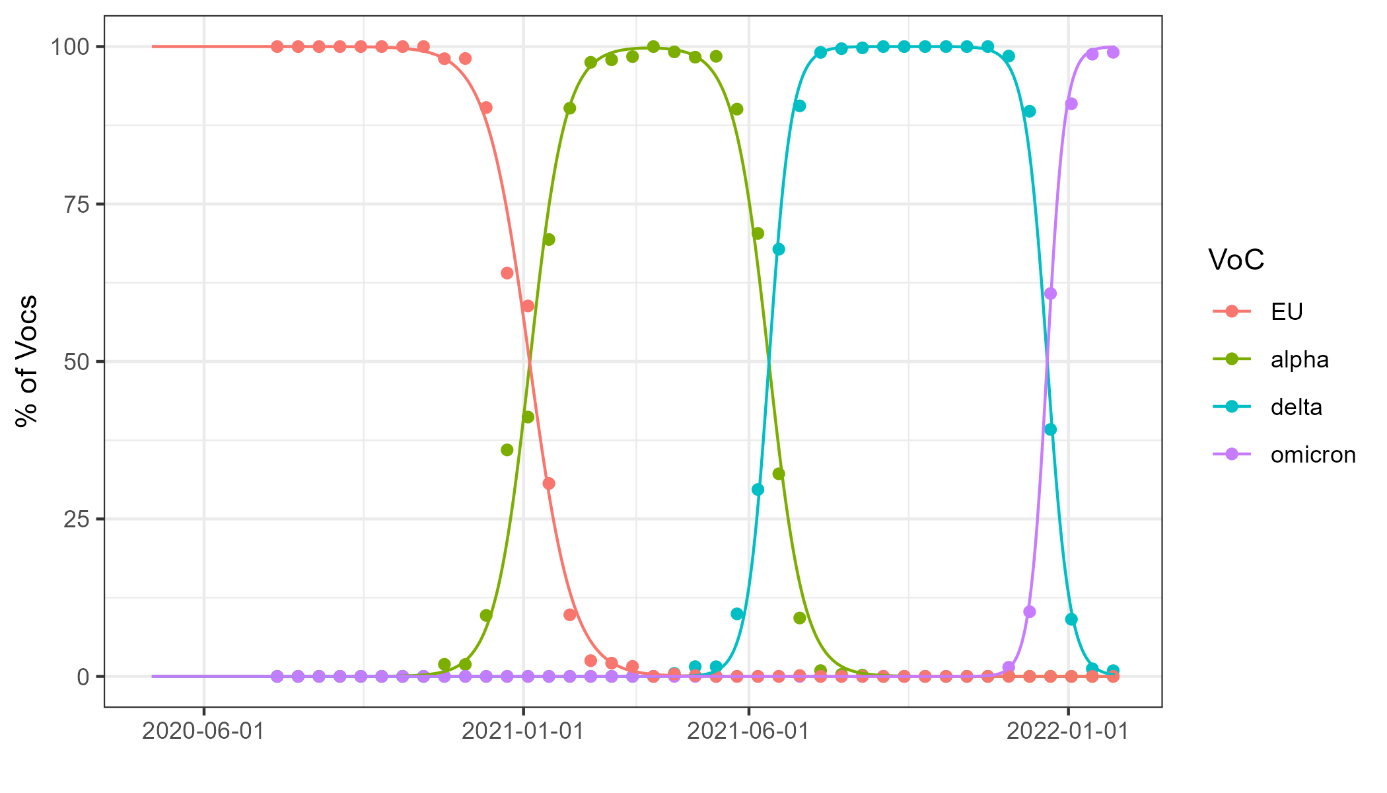

Supplementary figure S17: share of SarS-CoV-2 Variant of Concern (VoC) in the Geneva region: measures from GISEAD (points) and the estimation based on sigmoid functions (solid lines)

From this estimated curve, we determine the dates for two set of dominance threshold:
For 50% dominance:

| VoC | EU | alpha | delta | Omicron |
| --- | --- | --- | --- | --- |
| Start | 01-06-2020 | 06-01-2021 | 15-06-2021 | 18-12-2021 |
| end | 05-01-2021 | 14-06-2021 | 17-12-2021 | 01-02-2022 |

#### Non-pharmaceutical interventions

From the first COVID-19 positive case in Geneva (26-02-2020), several non-pharmaceutical interventions (NPI) have been put in place. The list and Figure S15 below summarize the main NPIs from March 2020 to February 2022:

- From 12-03-2020 to 26-06-2021: obligation to work from home
- From 16-03-2020 to 11-05-2020: confinement, including schools closure
- From 16-03-2020 to 30-05-2020: restriction of gathering to 5 people
- From 06-07-2020 to 26-06-2021: obligation to wear a mask in public
- From 21-10-2020 to 22-03-2021: restriction of gathering to 5 people
- From 02-11-2020 to 01-03-2021: closure of non-essential shops
- From 13-09-2021 to 17-02-2022: COVID certificate is mandatory in public places
- From 29-11-2021 to 17-02-2022: obligation to wear a mask in public places

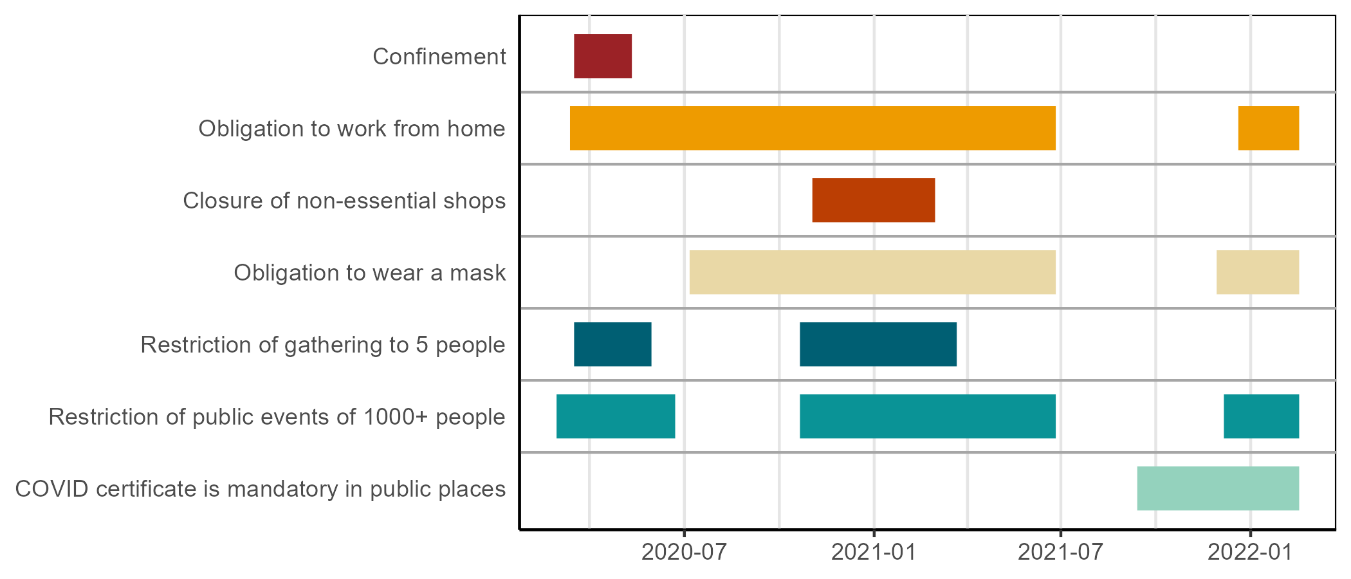

Fig. S15: main non-pharmaceutical interventions (NPI) in Geneva over time. The coloured lines indicate the period during which the NPIs described on the left were active.

During the complete period, both PCR and antigenic tests were free of charge for the population and there were no test accessibility restrictions except for a short period during the first EU1 epidemic wave. Antigenic tests were introduced the 09-11-2020. Note that the testing capacity was saturated during the peak period of the second EU1 wave (end of October 2020) and the Omicron wave (beginning of January 2022).

#### Vaccination

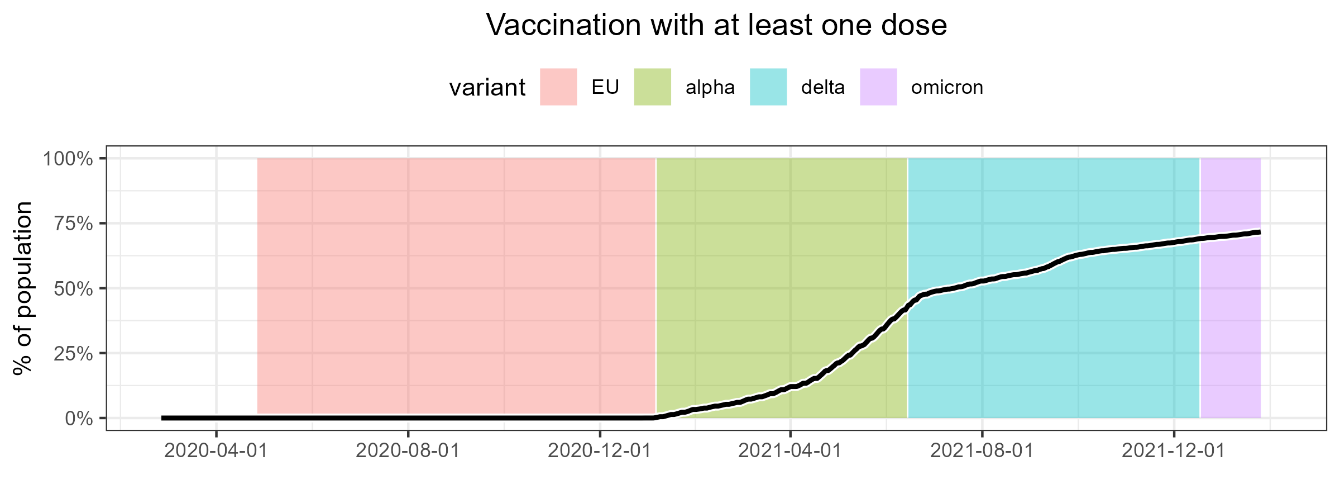

Fig. S16: Evolution in time of the proportion of the population with at least one dose in Geneva

.

In Geneva, the first vaccination centre opened the 25-01-2021 for person aged 75 or more at first. In February, vaccination was extended to vulnerable persons under 75 (immune-compromised persons, diabetic, …) and healthcare collaborators. The targeted population was progressively extended to people over 45 years old (from 12-04-2021), people over 16 years old (from 14-05-2021) and finally to children (from 02-07-2021 and from 12-01-2022 for younger people between 5 to 11 years old respectively). The most administrated vaccine type in Geneva was the RNA-based vaccine type, such as Moderna mRNA-1273 (59.83% of the total administrated vaccine doses in Geneva) and Pfizer BNT162b2 (39.86%), other type of vaccine stands for a minor part: Janssen (0.25%) and Nuvaxovid (0.06%). The final uptake at end of February 2022 was of 71.6% of the population who received at least one dose of vaccine (time evolution in Figure S16).

### Results

#### Influence of the delay

**Supplementary figure 1: Excess of concurrent infections at the addresses as a function of the delay between the two tests. The shaded area represent the 95% CI of the estimated number of secondary infections.**

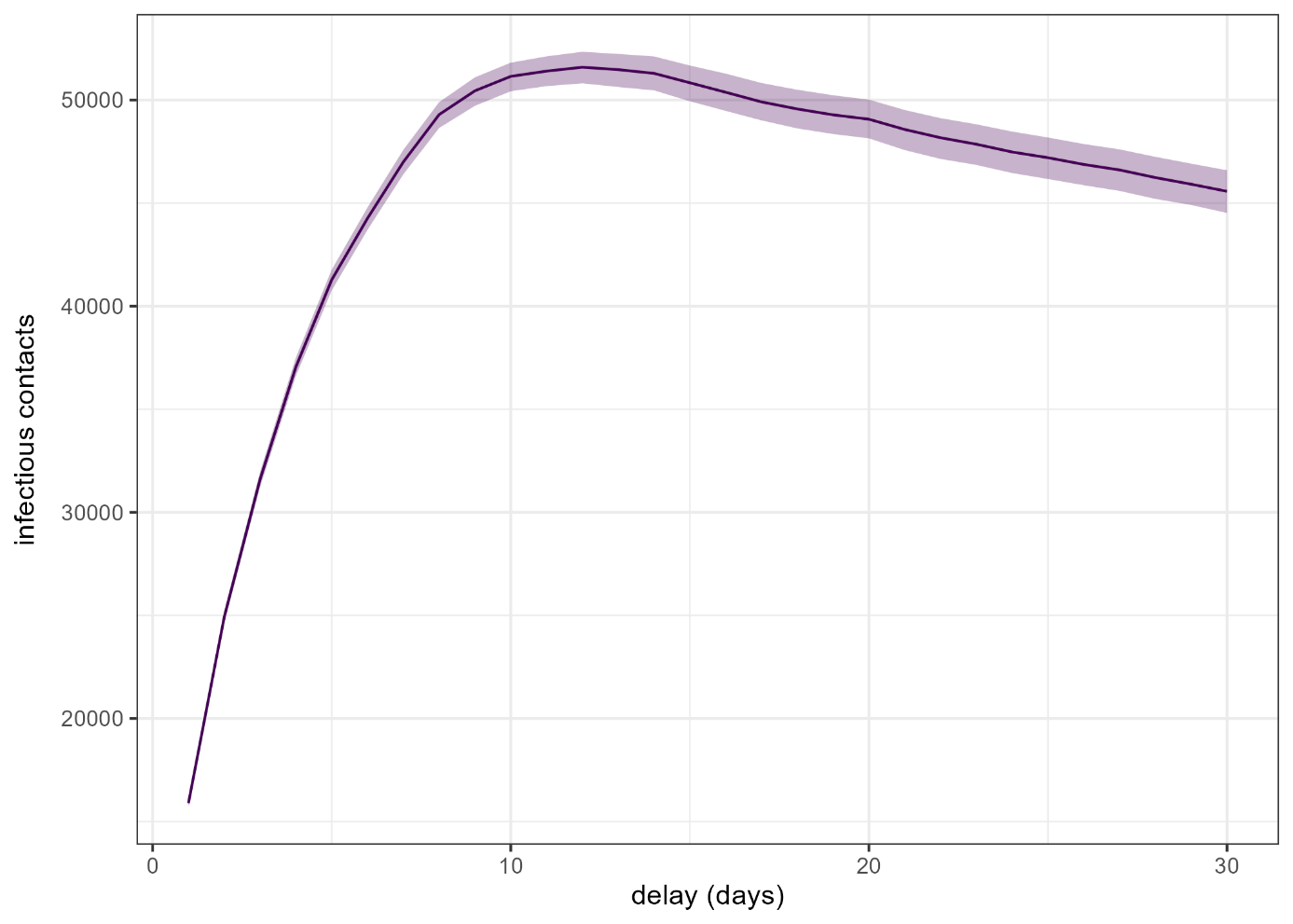

#### Sensitivity analysis

##### Delay of 6 days between tests

|  | **total** | **EU1** | **alpha** | **delta** | **omicron** |
| --- | --- | --- | --- | --- | --- |
| Number of raw concurrent infections | 123'304 | 28'574 | 7'297 | 14'888 | 72'545 |
| Number of concurrent infections in permutations | 78'429 CI: 79'443-77'445 | 15'337 CI: 15'583-15'104 | 1'702 CI: 1'784-1'630 | 6'140 CI: 6'298-5'998 | 55'250 CI: 55'778-54'713 |
| Estimated contagions at the address | 44'875 CI: 43'861-45'859 | 13'237 CI: 12'991-13'470 | 5'595 CI: 5'513-5'667 | 8'748 CI: 8'590-8'890 | 17'295 CI: 16'767-17'832 |
| Number of contacts declared living at the same address | 18'017 | 4'664 | 2'799 | 4'317 | 6'237 |
| percentage of contagions declared | 40.1% CI: 39.3-41.1 | 35.2% CI: 34.6-35.9 | 50% CI: 49.4-50.8 | 49.3% CI: 48.6-50.3 | 36.1% CI: 35-37.2 |

**Supplementary Table 1: Number of persons infected at the same address 6 days apart from each other, in the real data base, when permuting addresses on the state level (state base permutation), or on the neighbourhood level (neighbourhood base permutation). Estimations for permutation are the median of 1000 permutations given with their percentile confidence intervals (2.5% - 97.5% range).**

**Supplementary Table 2: Odds ratio (OR) with their associated confidence interval of the multivariable generalized model of under-reporting of the secondary infection, when considering concurrent infection as two positive tests 6 days apart. OR with and IR not encompassing 1 are grey shaded.**

| **Variable** | **EU1** | | **alpha** | | **delta** | | **omicron** | |
| --- | --- | --- | --- | --- | --- | --- | --- | --- |
|  | OR | 95% CI | OR | 95% CI | OR | 95% CI | OR | 95% CI |
| Contact age (reference: 17-65) |  |  |  |  |  |  |  |  |
| 0-16 | 1.36 | 0.93-1.9 | 1.24 | 0.91-1.65 | 1.16 | 0.93-1.44 | 1.13 | 0.93-1.34 |
| 65+ | 0.9 | 0.58-1.26 | 0.91 | 0.33-1.75 | 0.75 | 0.34-1.25 | 1.08 | 0.68-1.62 |
| Index age (reference: 17-65) |  |  |  |  |  |  |  |  |
| 0-16 | 1.19 | 0.75-1.7 | 1.29 | 0.92-1.83 | 1.04 | 0.84-1.29 | 1.3 | 1.08-1.55 |
| 65+ | 0.92 | 0.58-1.31 | 0.83 | 0.25-1.63 | 0.98 | 0.46-1.52 | 0.66 | 0.26-1.17 |
| Cati score (reference: 0) |  |  |  |  |  |  |  |  |
| 1 | 0.84 | 0.64-1.11 | 0.75 | 0.51-1.11 | 0.84 | 0.64-1.1 | 0.8 | 0.64-1 |
| 2-3 | 0.67 | 0.5-0.88 | 0.86 | 0.59-1.24 | 0.88 | 0.66-1.18 | 0.76 | 0.6-0.95 |
| 4-6 | 0.65 | 0.5-0.83 | 0.65 | 0.46-0.92 | 0.64 | 0.49-0.84 | 0.58 | 0.46-0.73 |
| Gender (reference: men-men) |  |  |  |  |  |  |  |  |
| Women-women | 1.22 | 0.81-1.81 | 1.09 | 0.72-1.62 | 1.34 | 0.98-1.82 | 1.05 | 0.82-1.37 |
| mixed | 1.27 | 0.91-1.81 | 1.23 | 0.89-1.78 | 1.24 | 0.94-1.64 | 1.06 | 0.85-1.36 |
| Type of building (single house) |  |  |  |  |  |  |  |  |
| House with isolated persons | 1.96 | 0.78-4.52 | 1.24 | 0.41-4.82 | 3.8 | 0.95-13.33 | 1.9 | 0.75-3.93 |
| Building without shops less than 40 inhabitants | 0.99 | 0.63-1.75 | 0.79 | 0.42-1.56 | 1.1 | 0.69-1.9 | 0.99 | 0.69-1.46 |
| Building without shops more than 40 inhabitants | 1.5 | 0.98-2.53 | 0.92 | 0.54-1.85 | 1.56 | 0.96-2.62 | 1.64 | 1.15-2.37 |
| Building with shops  less than 40 inhabitants | 1.57 | 0.99-2.6 | 1.17 | 0.61-2.45 | 1.78 | 1.06-3.18 | 2.06 | 1.44-3.13 |
| Building with shops  more than 40 inhabitants | 2.09 | 1.34-3.49 | 0.98 | 0.56-2 | 1.79 | 1.09-3.11 | 2.5 | 1.69-3.64 |
| Immune status (both non vaccinated) |  |  |  |  |  |  |  |  |
| Index : not vaccinated;  contact : vaccinated |  |  | 0.6 | 0.14-1.3 | 0.82 | 0.58-1.13 | 0.82 | 0.64-1.04 |
| Index : vaccinated;  contact : not vaccinated |  |  | 0.78 | 0.25-1.62 | 0.66 | 0.46-0.9 | 0.85 | 0.68-1.06 |
| Index : vaccinated;  contact : vaccinated |  |  | 1.78 | 0-10.53 | 1.04 | 0.69-1.52 | 0.9 | 0.7-1.12 |

##### Delay of 14 days between tests

**Supplementary Table 3: Number of persons infected at the same address 14 days apart from each other, in the real data base, when permuting addresses on the state level (state base permutation), or on the neighbourhood level (neighbourhood base permutation). Estimations for permutation are the median of 1000 permutations given with their percentile confidence intervals (2.5% - 97.5% range).**

|  | **total** | **EU1** | **alpha** | **delta** | **omicron** |
| --- | --- | --- | --- | --- | --- |
| Number of raw concurrent infections | 198'868 | 45'667 | 10'585 | 22'425 | 120'191 |
| Number of concurrent infections in permutations | 149'973 CI: 151'568-148'468 | 28'814 CI: 29'186-28'448 | 3'154 CI: 3'274-3'049 | 12'393 CI: 12'629-12'142 | 105'612 CI: 106'479-104'829 |
| Estimated contagions at the address | 48'895 CI: 47'300-50'400 | 16'853 CI: 16'481-17'219 | 7'431 CI: 7'311-7'536 | 10'032 CI: 9'796-10'283 | 14'579 CI: 13'712-15'362 |
| Number of contacts declared living at the same address | 21'733 | 5'592 | 3'821 | 5'244 | 7'076 |
| percentage of contagions declared | 44.4% CI: 43.1-45.9 | 33.2% CI: 32.5-33.9 | 51.4% CI: 50.7-52.3 | 52.3% CI: 51-53.5 | 48.5% CI: 46.1-51.6 |

**Supplementary Table 4: Odds ratio (OR) with their associated confidence interval of the multivariable generalized model of under-reporting of the secondary infection, when considering concurrent infection as two positive tests 14 days apart. OR with and IR not encompassing 1 are grey shaded.**

| **Variable** | **EU1** | | **alpha** | | **delta** | | **omicron** | |
| --- | --- | --- | --- | --- | --- | --- | --- | --- |
|  | OR | 95% CI | OR | 95% CI | OR | 95% CI | OR | 95% CI |
| Contact age (reference: 17-65) |  |  |  |  |  |  |  |  |
| 0-16 | 1.25 | 0.94-1.65 | 1.16 | 0.89-1.47 | 1.18 | 0.98-1.43 | 1.07 | 0.89-1.29 |
| 65+ | 0.93 | 0.61-1.27 | 0.91 | 0.44-1.61 | 0.91 | 0.52-1.41 | 1.11 | 0.68-1.65 |
| Index age (reference: 17-65) |  |  |  |  |  |  |  |  |
| 0-16 | 1.12 | 0.79-1.49 | 1.19 | 0.88-1.59 | 0.97 | 0.8-1.17 | 1.25 | 1.04-1.49 |
| 65+ | 0.93 | 0.64-1.3 | 0.89 | 0.41-1.55 | 0.94 | 0.53-1.4 | 0.71 | 0.3-1.2 |
| Cati score (reference: 0) |  |  |  |  |  |  |  |  |
| 1 | 0.88 | 0.7-1.13 | 0.77 | 0.56-1.06 | 0.91 | 0.7-1.16 | 0.82 | 0.65-1.01 |
| 2-3 | 0.68 | 0.52-0.88 | 0.84 | 0.61-1.13 | 0.91 | 0.7-1.16 | 0.78 | 0.62-0.96 |
| 4-6 | 0.64 | 0.51-0.8 | 0.63 | 0.47-0.83 | 0.66 | 0.52-0.86 | 0.59 | 0.47-0.72 |
| Gender (reference: men-men) |  |  |  |  |  |  |  |  |
| Women-women | 1.19 | 0.85-1.73 | 0.98 | 0.69-1.38 | 1.31 | 1.02-1.76 | 1.07 | 0.85-1.42 |
| mixed | 1.2 | 0.92-1.65 | 1.1 | 0.82-1.45 | 1.18 | 0.93-1.52 | 1.06 | 0.87-1.32 |
| Type of building (single house) |  |  |  |  |  |  |  |  |
| House with isolated persons | 1.95 | 0.83-3.99 | 1.69 | 0.54-5.09 | 3.4 | 1.15-8.98 | 2.5 | 1.07-4.72 |
| Building without shops less than 40 inhabitants | 1.2 | 0.78-2.04 | 0.76 | 0.44-1.42 | 1.16 | 0.78-1.84 | 1.31 | 0.92-1.9 |
| Building without shops more than 40 inhabitants | 2.08 | 1.37-3.48 | 1.11 | 0.65-1.95 | 1.87 | 1.31-2.89 | 2.33 | 1.67-3.34 |
| Building with shops  less than 40 inhabitants | 1.99 | 1.27-3.43 | 1.24 | 0.72-2.29 | 2 | 1.32-3.15 | 2.64 | 1.83-3.95 |
| Building with shops  more than 40 inhabitants | 2.94 | 1.85-5.03 | 1.18 | 0.7-2.15 | 2.27 | 1.49-3.53 | 3.6 | 2.54-5.22 |
| Immune status (both non vaccinated) |  |  |  |  |  |  |  |  |
| Index : not vaccinated;  contact : vaccinated |  |  | 0.67 | 0.21-1.37 | 0.85 | 0.65-1.11 | 0.85 | 0.67-1.08 |
| Index : vaccinated;  contact : not vaccinated |  |  | 0.7 | 0.27-1.36 | 0.72 | 0.52-0.96 | 0.85 | 0.68-1.04 |
| Index : vaccinated;  contact : vaccinated |  |  | 1.45 | 0-5.94 | 0.94 | 0.67-1.31 | 0.9 | 0.72-1.13 |
